# Accurate, Robust, and Scalable Machine Abstraction of Mayo Endoscopic Subscores from Colonoscopy Reports

**DOI:** 10.1101/2022.06.19.22276606

**Authors:** Anna L. Silverman, Balu Bhasuran, Arman Mosenia, Fatema Yasini, Saransh Gupta, Taline Mardirossian, Rohan Narain, Justin Sewell, Atul J. Butte, Vivek A. Rudrapatna

**Affiliations:** Division of Gastroenterology and Hepatology, Department of Medicine, Mayo Clinic, Phoenix, AZ, 85054; Department of Medicine, University of California, San Diego, La Jolla, CA, 92093; Bakar Computational Health Sciences Institute, San Francisco, CA, 94143; School of Medicine, University of California San Francisco, San Francisco, CA 94143; Department of Computer Science, University of California Berkeley, Berkeley, CA, 94720; Division of Gastroenterology, Department of Medicine, Zuckerberg San Francisco General Hospital, San Francisco, CA, 94110; Center for Data-Driven Insights and Innovation, University of California Health, Oakland, CA, 94607; Division of Gastroenterology and Hepatology, Department of Medicine, University of California, San Francisco, San Francisco, CA, 94143

## Abstract

**Importance:** Electronic health records (EHR) data are growing in importance as a source of evidence on real-world treatment effects. However, many clinical important measures are not directly captured as structured data by these systems, limiting their utility for research and quality improvement. Although this information can usually be manually abstracted from clinical notes, this process is expensive and subject to variability. Natural language processing (NLP) is a scalable alternative but has historically been subject to multiple limitations including insufficient accuracy, data hunger, technical complexity, poor generalizability, algorithmic unfairness, and an outsized carbon footprint.

**Objective:** Compare different algorithmic approaches for classifying colonoscopy reports according to their ulcerative colitis Mayo endoscopic subscores

**Design:** Other observational study – NLP algorithm development and validation

**Setting:** Academic medical center (UCSF) and safety-net hospital (ZSFG) in California

**Participants:** Patients with ulcerative colitis

**Exposures:** Colonoscopy

**Main Outcomes and Measures:** The primary outcome was accuracy in identifying reports suitable for Mayo subscoring (binary yes/no) and then separately assigning a Mayo subscore where relevant (ordinal). Secondary outcomes included learning efficiency from training data, generalizability, computational costs, fairness, and sustainability.

**Results:** Using automated machine learning (autoML) we trained a pair of classifiers that were 98% [91-99%] accurate at determining which reports to score and 97% [88-99%] accurate at assigning the correct Mayo endoscopic subscore. The binary classifiers trained on UCSF data achieved 96% accuracy on hold-out test data from ZSFG. Training these classifiers required 4 hours of computation on a standard laptop. Classification errors were not associated with either gender or area deprivation index. The carbon footprint of this approach was 24x less than current deep learning algorithms for clinical text classification.

**Conclusions and Relevance:** We identified autoML as an efficient and robust method for training clinical text classifiers. AutoML-trained classifiers demonstrated many favorable properties including generalizability, limited effort needed for data annotation and algorithm training, fairness, and sustainability. More generally, these results support the feasibility of using unstructured EHR data to generate real-world evidence and drive continuous improvements in learning health systems.

**Key Points:** *Question:* Is natural language processing (NLP) a viable alternative to manually abstracting disease activity from procedure notes?

*Findings:* We compared different methods for abstracting the ulcerative colitis Mayo endoscopic subscore from colonoscopy reports. Classifiers trained using automated machine learning (autoML) achieved the greatest accuracy (97%), recognized when to abstain, generalized well to other health systems, required limited effort for annotation and programming, demonstrated fairness, and had a small carbon footprint.

*Meaning:* NLP methods like autoML appear to be sufficiently mature technologies for clinical text classification, and thus are poised to enable many downstream endeavors using electronic health records data.

## Introduction

Real-world evidence (RWE) refers to the use of observational data for the study of diseases and treatments.^1,2^ Interest in using electronic health records (EHRs) for RWE continues to grow, as these systems capture detailed data on patient diagnoses, treatments and outcomes over time. As such, they have the potential to be useful for many purposes, from regulatory evaluations of drugs and devices to scalable assessments of healthcare quality and value.^3,4^

A challenge with using these data is that many critical elements are captured in a free-text form rather than analysis-ready data. Thus, many EHR-based analyses utilize the structured data alone and ignore the content in notes. Although this approach is attractive because of its feasibility, it increases the risks of confounding and other biases in observational research.^5,6^ This is particularly the case for chronic conditions where disease assessments are complex and captured as free-text, and where these assessments inform decision making. Manual abstraction of these elements from notes has been a common strategy to minimize this bias. However, this approach is fundamentally unscalable. It also requires expertise and is subject to interrater variability and reviewer fatigue.

Natural language processing (NLP) refers to computational methods for analyzing language-related data. The use of NLP on clinical text has been an active field for several decades, with dozens of software packages now freely available.^7-10^ While these technologies have opened opportunities for scalable and robust analyses of real-world data, they too are not without their challenges. These methods have generally not been sufficiently developed or validated for most abstraction tasks due to their sheer number. Thus, dedicated efforts by individual groups are typically needed to evaluate and enhance these methods for specific uses. Doing this can require significant technical capabilities, including programming experience and specialized hardware. Many modern NLP methods can be very data inefficient, placing large burdens on annotators and decreasing the overall scalability of the algorithm-development process.

Lastly, these methods can be associated with many negative societal consequences. Many black box methods can overfit on clinically immaterial features like socioeconomic status. As a result, they can propagate healthcare disparities and threaten algorithmic robustness unless dedicated precautions are taken.^11-14^ Algorithmic training can also be unsustainable for the environment, with single algorithms being associated with a carbon footprint equivalent to the lifetime emission of five cars.^15^

We sought to determine if current NLP methods can be considered a viable alternative to manual abstraction from the EHR. Here we report the results of a comprehensive and comparative assessment of several methods for training text classifiers, designed to assess their current utility for RWE studies. We selected a use case of ulcerative colitis (UC), a chronic inflammatory disease of the large intestine. Using the colonoscopy reports from two medical centers, we compared different classifiers by their ability to process colonoscopy reports and abstract the Mayo endoscopic subscore, a disease activity measure commonly used in UC registrational trials.^16^ Our primary endpoint was accuracy on the sequential tasks of identifying which reports could be scored using the Mayo endoscopic subscore and assigning a subscore if appropriate. Secondary endpoints included learning efficiency, generalizability, programming effort, algorithmic fairness, and carbon emissions.

## Methods

### Procedure Reports

To identify colonoscopy reports for classifier training and evaluation, we accessed the EHRs at two health systems in California: an academic medical center (University of California, San Francisco; UCSF) and a safety-net hospital (Zuckerberg San Francisco General; ZSFG). These institutions have different physician groups and use different endoscopy reporting software. We queried the back end EHR databases to identify all patients who had ever been assigned an ICD-10 code for inflammatory bowel disease (K50*, K51*), and extracted all corresponding colonoscopy reports from the 2017-2020 period.

### Annotation Procedure

In the first stage of this procedure, two physicians uniformly sampled and annotated reports as being suitable for Mayo subscoring or not. The main criteria for defining suitability included a clear diagnosis of UC and surgically unaltered anatomy (Supplemental Methods). This was recorded as a binary variable. This procedure continued until at least 75 eligible reports per site and per annotator were annotated.

In the second stage, suitable reports were assigned a Mayo endoscopic subscore, an ordinal measure of UC disease activity that ranges from 0 through 3. Scores were assigned based on the most severely affected segment, and with any friability scored as a 2 or higher (Supplemental Methods).^16^

The interrater agreement of this process was assessed on a separate set of 50 uniformly sampled reports.

### Algorithm Development and Validation

We developed and evaluated four standard methods for abstracting information from notes: cTAKES^7^-based concept recognition, bag-of-words models using sklearn^8^ and automated machine learning (autoML^9^), as well as three models related to BERT^10^ (Supplemental Methods). These methods vary in their underlying technique, requirements for training data, tendencies towards robust and generalizable learning, and ease of use.

We used these methods to separately train a binary classifier (to predict which procedure reports were Mayo scorable) and an ordinal classifier (to predict the correct Mayo endoscopic subscore for scorable reports). As a control, we developed null classifiers that predict the dominant class for each task. All classifiers were evaluated on a 20% held-out test set stratified by score, annotator, and site.

The classifier achieving the highest accuracy was subjected to additional evaluations of generalizability and learning efficiency. To assess generalizability, we retrained the binary classifier on the data from UCSF alone and evaluated it on data from ZSFG. There were insufficient reports to adequately assess generalizability for the ordinal prediction task due to the multiplicity of classes and class imbalance.

### Algorithmic Fairness

We evaluated the fairness of our algorithms by estimating their misclassification rate along lines of gender and social deprivation. We accessed patient-level structured data at UCSF to perform these analyses. We used the area deprivation index (ADI) mapped to residential zip codes as a proxy for social deprivation.^17^ Sex assigned at birth was unavailable in our database, and we were unable to perform analyses by race or ethnicity due to insufficient procedure notes from each race/ethnicity.

### Carbon Emissions

We used a public machine learning (ML) emission calculator to estimate the environmental impact of our algorithms (Supplemental Methods).^18^

### Statistics

We computed exact binomial confidence intervals for all results reported as a sample proportion. For analyses of algorithmic fairness, we used ordinal logistic regression to separately model the misclassification error as a function of either gender or ADI. We report the corresponding p-values from a Wald test. We performed all computing using *R 4*.*1*.*3* and *Python 3*.

### Ethics

The study was approved by the UCSF Institutional Review Board (#18-24588).

## Results

### Procedure Reports

The source corpus consisted of 3,769 notes from UCSF and 835 from ZSFG, all authored between 2017 and 2020 (Figure 1). The manually annotated corpus consisted of 499 notes of which 282 were from UCSF and 217 from ZSFG. 302 notes were eligible for Mayo endoscopic scoring, with 151 notes from each site. Inter-annotator agreement was 96% [86-100%] for the binary task (N=50) and 88% [69-97%] for the ordinal task (N=25).

**Figure 1:**
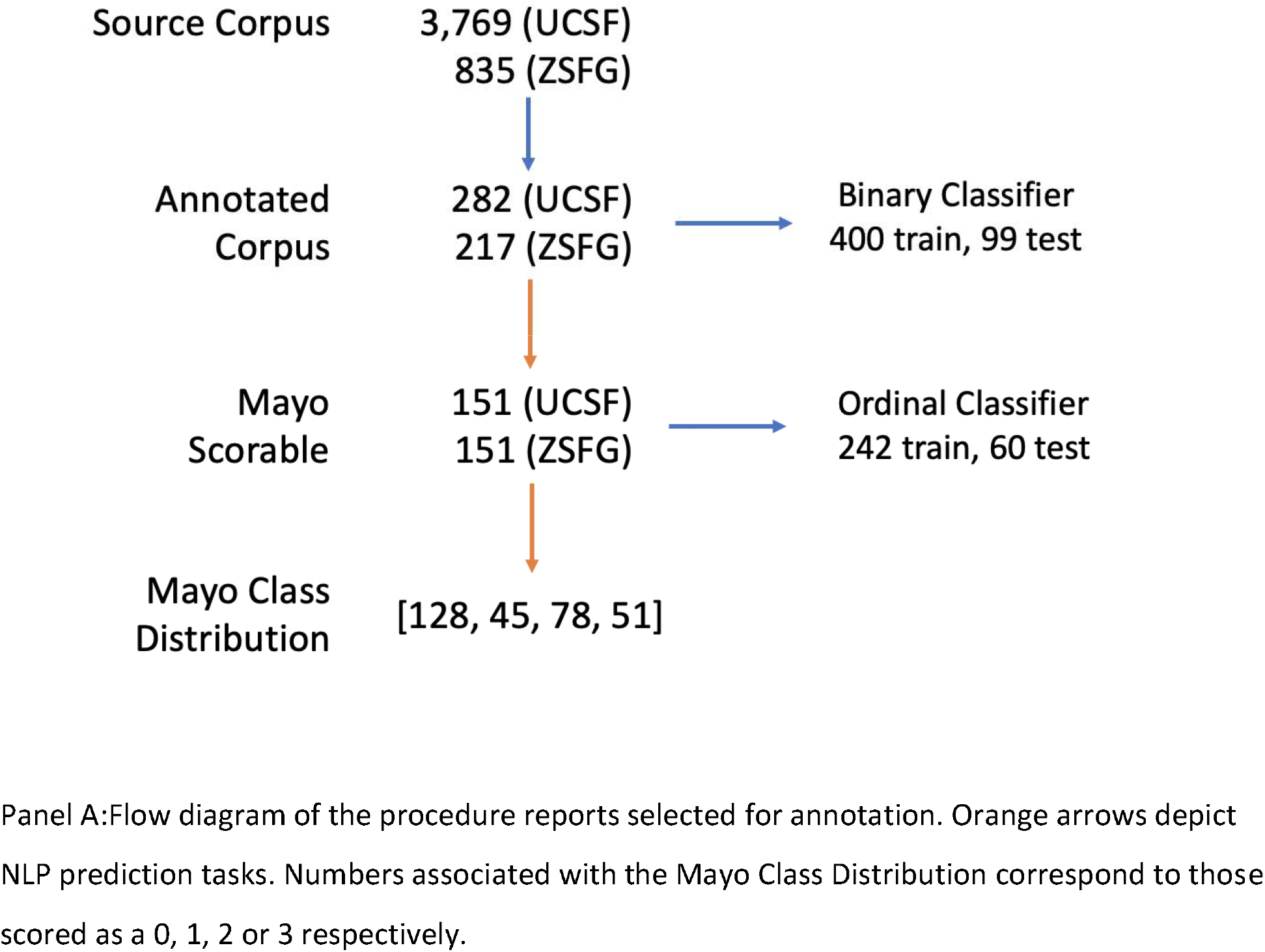
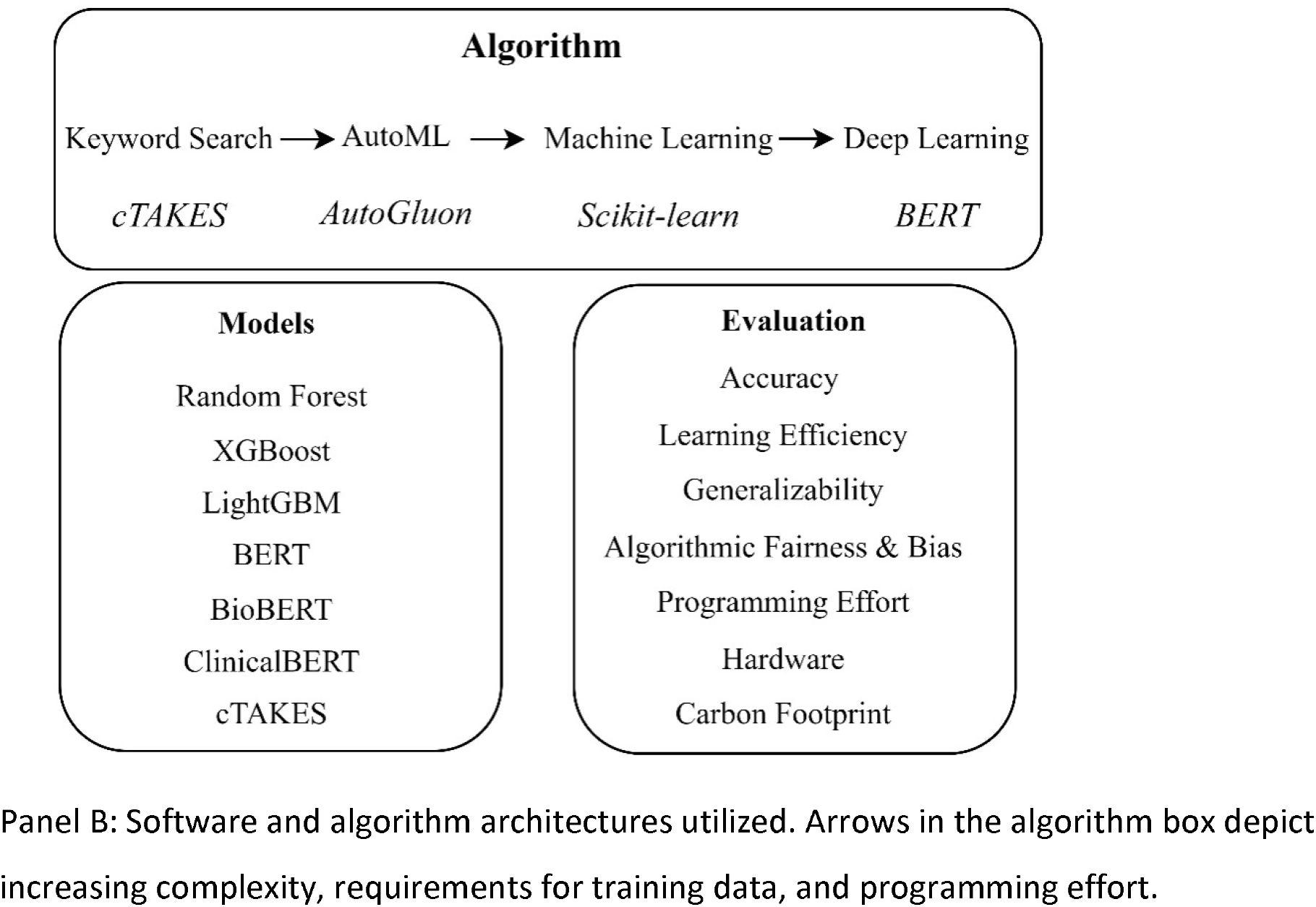
Corpus Flow Diagram and Algorithm Architectures.

### Algorithmic Accuracy

The autoML-trained classifiers achieved the overall highest accuracy. They were 98% [91-99%] accurate at identifying Mayo scorable reports and 97% [88-99%] accurate at assigning a Mayo subscore (Table 2). The relative ordering of algorithmic performance was preserved across both tasks, with the sklearn classifiers consistently outperforming the BERT-based classifiers. ClinicalBERT was substantially more performant than BioBERT and BERT-base, presumably reflecting the sensitivity of these algorithms to pretraining data: ClinicalBERT was trained on clinical notes, whereas BioBERT and BERT-base were trained on biomedical journal articles and general Wikipedia articles respectively. To our surprise our manually designed, rule-based approach utilizing cTAKES-recognized clinical concepts was the least accurate (22%). It performed worse than a null model that predicts the most common subscore for all reports (42%).

**Table 1:** Mayo Endoscopic Score Keywords. Keywords used to delineate each Mayo score for the keyword search-based algorithm. All procedure reports were classified according to the highest Mayo score for which a corresponding, non-negated concept was identified.

| Mayo Score | Keywords |
| --- | --- |
| 0 | 'normal', 'quiescent', 'scar' |
| 1 | 'erythema', 'decreased vascular pattern', 'granularity', 'aphthous ulcer', 'aphthae', 'mild' |
| 2 | 'friability', marked or extensive 'erythema', 'loss of vascularity', 'absent vascularity', 'erosions', 'moderate' |
| 3 | spontaneous 'bleeding', 'ulcer', 'ulcerated', 'severe' |

**Table 2:** Algorithmic performance for the Mayo scorability (Binary) and the Mayo endoscopic subscore (Ordinal) prediction tasks.

| Algorithm | Framework | Type | Accuracy (95% CI) | F-Score | Programming Effort | Lines of Code | Hardware |
| --- | --- | --- | --- | --- | --- | --- | --- |
| RandomForest | autoML | Binary | **98% (91-99)** | **97.47** | 1 week | 12 | CPU |
| LightGBM | Scikit-learn | Binary | **98% (89-98)** | 97.00 | 4 days | 124 | CPU |
| ClinicalBERT | Transformer | Binary | 93% (84-96) | 93.00 | 3 weeks | 146 | GPU |
| Null Model | Scikit-learn | Binary | 60% (50-70) | - | - | - | - |
| BERT | Transformer | Binary | 47% (37-57) | 44.68 | 3 weeks | 146 | GPU |
| BioBERT | Transformer | Binary | 38% (28-48) | 39.00 | 3 weeks | 146 | GPU |
| XGBoost | autoML | Ordinal | **97% (88-99)** | **97.00** | 1 week | 12 | CPU |
| LightGBM | Scikit-learn | Ordinal | 77% (63-87) | 77.10 | 4 days | 124 | CPU |
| ClinicalBERT | Transformer | Ordinal | 61% (48-74) | 61.66 | 3 weeks | 146 | GPU |
| Null Model | Scikit-learn | Ordinal | 41% (29-55) | - | - | - | - |
| BioBERT | Transformer | Ordinal | 50% (37-63) | 50.00 | 3 weeks | 146 | GPU |
| BERT | Transformer | Ordinal | 38% (26-52) | 38.33 | 3 weeks | 146 | GPU |
| Rule based using clinical concepts | cTAKES, RegEx | Ordinal | 22% (12-37) | 22.10 | 1 day | 19 | CPU |

### Learning efficiency, technical feasibility

Although high accuracy is typically the primary objective of most endeavors involving NLP, practicality is an almost equally important consideration. Many supervised learners like BERT can require hundreds to thousands of expert annotations, an unrealistic requirement in many contexts. We measured the learning efficiency of the autoML classifiers by measuring their performance using decreasing subsets of the training data. The binary classifier remained 95% [82-96%] accurate despite training on only 240 notes (60% of the training data) (Table 3). On the ordinal task, the accuracy dropped from 97% [88-99%] to 70% [66-74%] when reducing the training data from 242 notes to 194 notes (80% of the dataset). This was consistent with our expectation that more data would be required for the ordinal task, given the multiplicity of target classes and degree of class imbalance. These results supported the utility of autoML as a method for efficiently developing text classifiers in practical contexts.

**Table 3:**
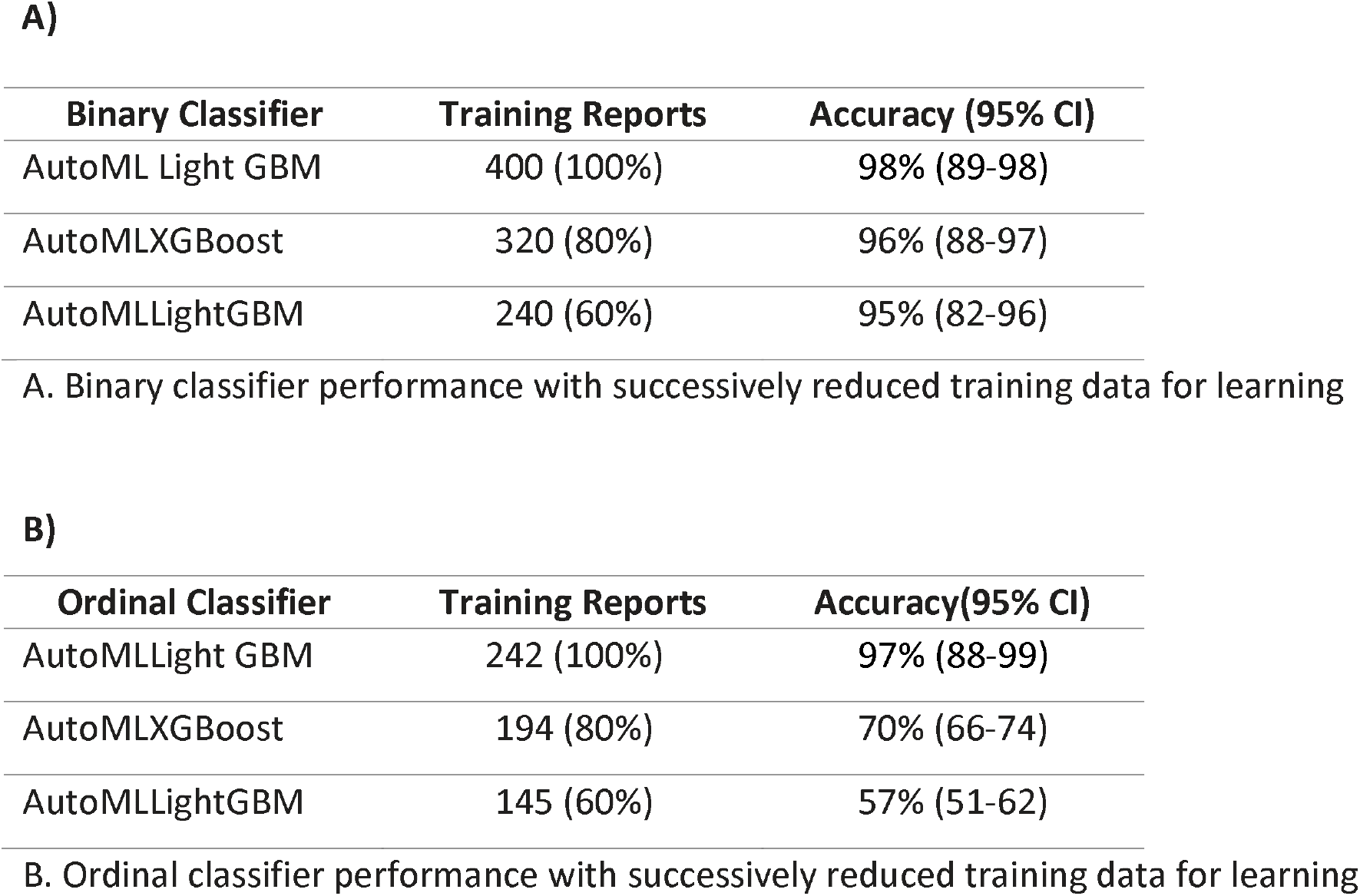
Learning efficiency of AutoML Algorithms

AutoML appeared favorable from a technical perspective as well. We produced these classifiers with 4 hours of computation on a standard laptop. By contrast, the BERT-related models required substantial technical troubleshooting on a hardware-intensive environment.

### Generalizability

Many machine learning models are prone to overfitting on irrelevant predictive features and thus fail to generalize to data from other health systems. We assessed the robustness of the finalized autoML classifiers by re-training them on just the UCSF data and evaluating it on data from ZSFG. On the binary prediction task, classifiers trained on the 282 UCSF reports remained 96% accurate when evaluated on the 217 ZSFG notes. We did not assess the ordinal classifier due to insufficient data (only 151 available reports) considering the learning efficiency results as reported above.

### Social impacts

There has been a growing awareness of the impacts that artificial intelligence has on society in recent years. For example, the rise of automation and our trust in it has the potential to propagate existing social disparities, a phenomenon known as algorithmic unfairness. In addition, the training of some models such as BERT can be surprisingly unsustainable for the environment, with carbon footprints equivalent to the lifetime exhaust produced from five cars.^15^ We assessed our automML classifiers along these two dimensions: fairness and sustainability.

We used linked EHR data to map procedure reports to patient gender, and mapped area deprivation index (ADI) via residential zip code. We found no evidence of bias by either of these factors, with p-values of 0.65 and 0.80 respectively. We could not assess other variables of *a priori* importance like race and ethnicity due to severe class imbalance.

Finally, we used a previously published method for quantifying the carbon emissions of machine learning algorithms.^18^ AutoML training required 0.0216 kilogram equivalents of CO_2_, comparable to charging a smartphone 3 times.^19^ This was 24 times less costly than the process of fine tuning ClinicalBERT on the annotated dataset, and roughly 5,000 times less costly than training a dedicated, BERT-base algorithm.

## Discussion

We compared several computational methods for transforming routinely documented clinical text into quantitative, analysis-ready data. As our use case we selected two sequential tasks related to the Mayo endoscopic subscore, a key measure of ulcerative colitis disease activity. The method yielding the best results across a wide range of metrics was automated machine learning (autoML), a computationally lean and powerful framework for training supervised learning models. These classifiers were highly accurate at assigning Mayo subscores to colonoscopy reports and recognizing when to abstain. They appeared to learn robust and generalizable predictive features while requiring only a limited amount of effort for annotations and programming. They outperformed BERT-based classifiers, which hold state of the art status on many NLP tasks, as well as cTAKES, a well-established software suite for clinical NLP. Lastly, they demonstrated evidence of algorithmic fairness and environmental sustainability.

Recent years have seen a growing interest in the use of electronic health records data to close evidence gaps and improve quality and value in healthcare. However, a persistent bottleneck in the optimal use of these data has been the large burden of analytically inaccessible data captured as free text. Over the past several decades, clinical NLP has made substantial strides towards the goal of accurate and practical computational alternatives to manual data abstraction. It has explored a multitude of technical approaches, from rule-based expert systems incorporating clinical knowledge to data-driven, supervised machine learning. Progress in this field has generally been uneven, with different solutions typically reflecting tradeoffs between accuracy, flexibility, generalizability, learning efficiency, and technical expertise. Consequently, the use of NLP in clinical research and operations has largely remained in the domain of specialized technical laboratories but otherwise failed to achieve widespread adoption to date.

Our results provide early evidence that the field of “applied clinical NLP” has finally arrived. It appears that we finally have enough tools at our disposal to be able to solve many clinical text-related tasks in a scalable way, and perhaps NLP can realistically be considered as a first-line option for many clinical teams with limited resources and technical expertise. The advent of autoML technologies exemplifies this perspective particularly well, given their unique combination of performance and user friendliness.

However, we note that no algorithm will solve all problems equally well, and different solutions will be required for different types of problems.^20^ We outline our approach in Table 4. Simple tasks that may not require textual context may benefit from keyword- and rule-based methods, whereas complicated classification tasks may require supervised approaches like autoML and BERT. It is worth noting that it is not always easy to characterize the difficulty of tasks *a priori*. We expected the rule-based model to perform well because of the clear mapping between natural language descriptors and Mayo subscores. The reasons for its mediocre performance remain unclear. We elected not to investigate this further given the identification of far more performant models and the overall objectives of this study.

**Table 4:**
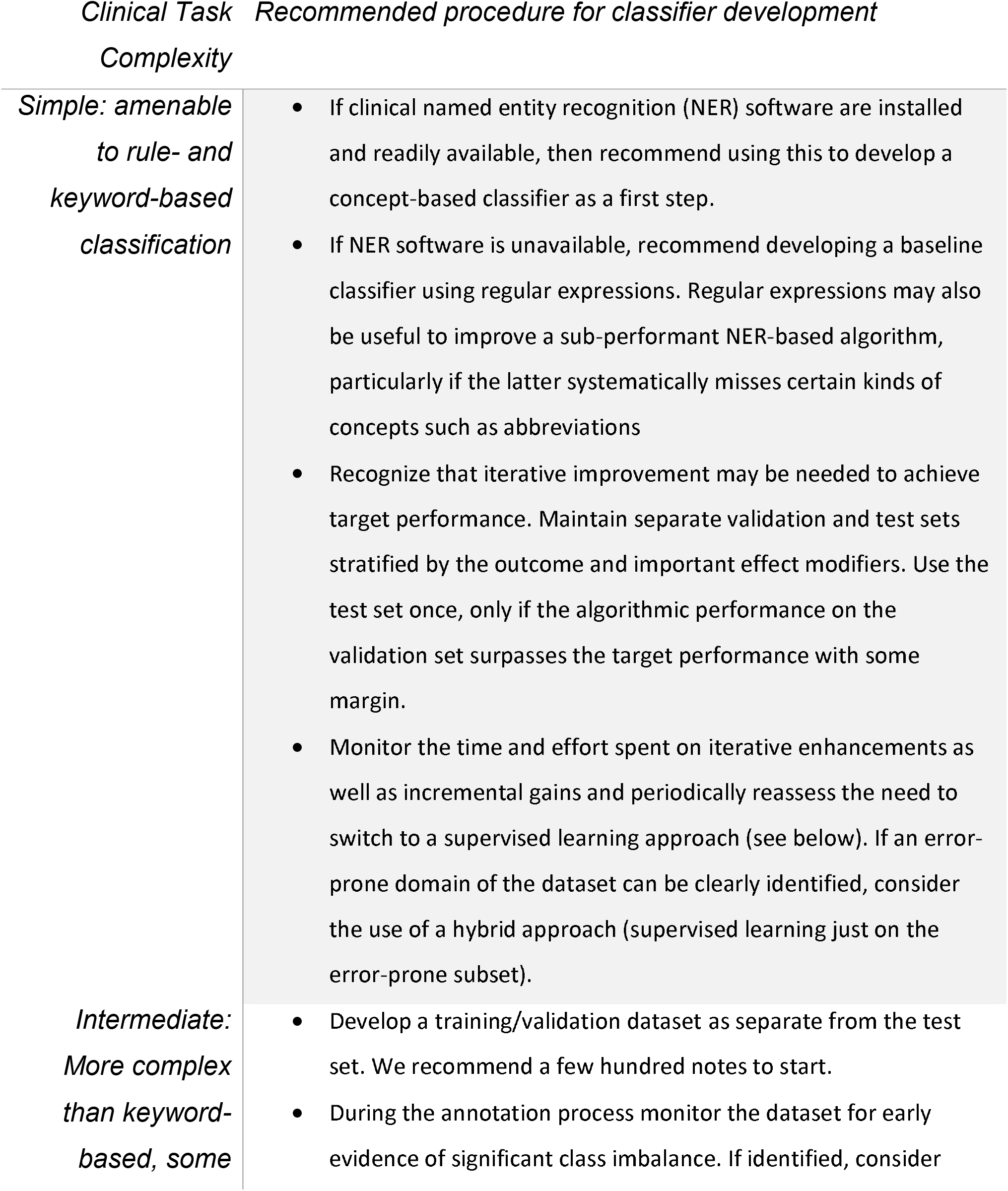
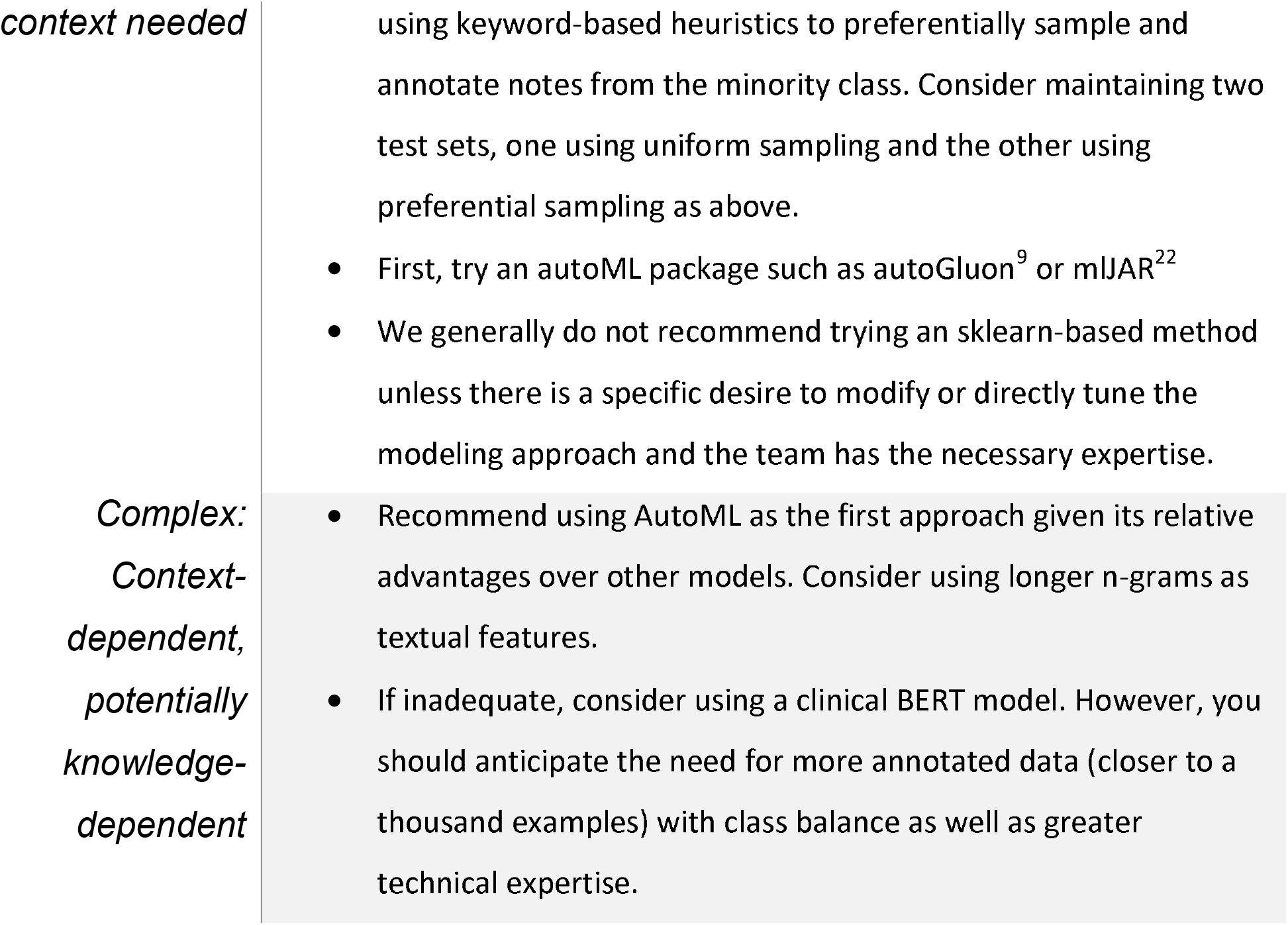
Selecting the best algorithmic approach for clinical information extraction and/or text classification

Another issue that deserves mention is class imbalance, a common feature in real-world data. In our study, there was a significant class imbalance for the Mayo 3 reports. We suspect that the suboptimal performance of the BERT models for the ordinal classification task was precisely for this reason. By contrast, ClinicalBERT performed extremely well on the class-balanced, binary classification task. Future solutions to this problem could include preferential sampling with keyword searches, to selectively annotate notes from the minority class.

Algorithms, especially those that rely on gold-standard annotations such as ours, are susceptible to the influence of bias similar to humans.^11^ We tested and found no association between our classifier’s misclassification rates and either gender or ADI, a measure of socioeconomic status. However, we acknowledge that our study was insufficiently powered to exclude important degrees of bias. Moreover, we note that our assessment only evaluated algorithmic fairness relative to the gold-standard produced by the annotators. This study design cannot exclude possible bias by the original clinician who documented the procedure report. Nonetheless we propose that all algorithms, especially those applied to healthcare, undergo formal evaluations for possible bias. Future work is needed to establish standards of fairness and ensure that unchecked algorithmic biases do not propagate at scale.

Finally, we note that our autoML classifiers were associated with a small carbon footprint. The emissions associated with these models was 24 times less than a fine-tuned ClinicalBERT, and roughly 5,000 times less than a dedicated BERT model. We do acknowledge that the relative environmental stakes between the choice of an autoML model versus a pre-trained BERT model are modest, considering the innumerable and substantial sources of anthropogenic carbon emissions. However, we foresee the continued growth of machine learning and data science across a range of endeavors including healthcare. It is incumbent upon all of us using these technologies to do our part, raise awareness within our communities of practices, and curb these multiplicative effects.^21^ We must also modernize our reporting standards, and treat the reporting of environmental self-audits with the same level of rigor as we have come to expect with other ethics-related disclosures in research.

Our study has several strengths. We utilized a multicenter corpus of procedure notes encompassing differences in physicians and their documentation styles, patients, and procedure reporting software. We ensured acceptable agreement between expert annotators. In addition to typical metrics like accuracy, we paid attention to feasibility and other barriers to widespread adoption, including data hunger and technical requirements. We assessed the social impacts of our models, including algorithmic fairness and carbon footprint. We are releasing the analytic code to reproduce and extend these results for other uses.

We acknowledge several limitations. Some of our analyses were underpowered or lacked sufficient data to be analyzed, such as the assessments of algorithmic fairness. Our test set was relatively small. Although we followed standard model development procedures to limit overfitting and employed stratified sampling of the test set, we cannot rule out the possibility of some residual bias from the algorithm procedure itself. Nonetheless, we believe that our primary results pertaining to the rank-ordering of algorithms are likely to remain robust to this. We noted that the accuracy of the ordinal classifier (97%) was greater than the point estimate of interrater reliability on this task (88%).However the 95% confidence intervals are consistent with statistical equivalence, and the performance of these models on the generalizability assessment suggest that any overfitting is likely small. Finally, we cannot comment on the degree to which these findings will apply to a broader range of real-world tasks. More work is needed to investigate the generalizability of our findings.

In conclusion, we conducted a multicenter assessment of computational methods for performing text classification. We adopted a use case of abstracting Mayo endoscopic scores from colonoscopy procedure reports. We found that classifiers trained using autoML performed well across a range of metrics, including accuracy, generalizability, learning efficiency, programming effort, technical requirements, fairness, and carbon footprint. We propose that this method be a key element in the toolkit of clinical and data science teams working with free-text, and that this overall technology be considered a viable option for situations where efficient, scalable, and consistent annotations of text are needed.

## Data Availability

The analytic code has been made publicly available at https://github.com/rwelab/MayoClassifier. The data used for this study contains protected health information and thus have not been made available for reuse. However, a machine-redacted version of the data can be made available to requesting researchers by mutual agreement and following the execution of a data use agreement.

## Previous Presentation of Information

Prior versions of this work have been presented at the American College of Gastroenterology Meeting 2020 and Digestive Diseases Week 2022.

