## Supplemental Methods for "Accurate, Robust, and Scalable Machine Abstraction of Mayo Endoscopic Subscores from Colonoscopy Reports"

**Algorithm Development and Validation**

We developed and evaluated four standard methods for abstracting information from clinical notes. These methods vary in their underlying technique (rule-based vs supervised learning), requirements for training data, tendencies towards robust and generalizable learning, and ease of use.

Our first approach was a rule-based classifier. First, we used clinical named entity recognition software (cTAKES^7^ version 3.1.1) and regular expressions (RegEx) to identify relevant clinical concepts and concept negation. Then, we manually defined a rule-based algorithm to assign a Mayo endoscopic subscore based on the most severe descriptor of disease activity. Reports that contained at least one concept were classified according to the highest corresponding subscore. Reports without any identifiable concepts were classified as a 0.We did not use this approach to generate a classifier for Mayo scorability, given the heterogeneity of non-Mayo scorable reports and the difficulty in manually specifying an algorithm.

We used supervised machine learning (ML) to train all subsequent classifiers. Our second approach utilized scikit-learn^8^ (sklearn), a popular framework for training and evaluating ML models in Python. We trained 30 different bag-of-words classifiers using unigram, bigram and trigram-based predictors and default hyperparameters. For sklearn-based classifiers, we trained multiple linear and tree-based algorithms including Naive Bayes, KNeighbors, Logistic Regression, Decision Tree, Random Forest, Extra Tree, Support vector machines (SVMs), AdaBoost (Adaptive Boosting), XGBoost (eXtreme Gradient Boosting) and LightGBM (Light Gradient Boosting Machine). We used uni, bi, and tri-gram features, and performed vectorization separately for the train and test set to avoid information leakage. Specifically, we generated the training set matrix using the vectorizer.fit_transform function and generated the test set matrix using the vectorizer.transform function. We trained all of these classifiers using the default hyperparameter settings in scikit-learn 1.0.2.

We trained a third set of classifiers using automated machine learning (autoML), an end-to-end approach that streamlines many aspects of classifier development such as feature engineering, hyperparameter tuning, and ensembling.^23^ Using the same n-gram features as above, we trained 75 different classifiers which included Random Forest and XGBoost. We used the open-source package *autoGluon**^9^*to train these classifiers.For the autoML classifiers we used *AutoGluon* version 0.3.1. The *AutoGluon* pipeline includes data preprocessing (e.g. prediction problem inference, label mapping), feature generation (e.g. ngram, astype, fillna, identity, category, dropunique, continuous vs discrete, rescaling) model fitting, evaluation, and inference. We generated models namely Logistic Regression, Random Forest, ExtraTrees, KNeighbors, LightGBM, XGBoost, CatBoost ,NeuralNetFastAI, Weighted Ensemble, StackerEnsemble, Bagged Ensemble. Additioanal models were generated using stack level (x) the model is trained in such as “_L1”, “_L2”, etc. (eg: LightGBM_BAG_L1, LightGBM_BAG_L2), hyperparameter search (HPO) (eg: LightGBM/T4), and bagged ensemble (eg: LightGBM_BAG_L1). The generated models include baseline classifiers as well as algorithm ensembles using stacking and bagging. We developed these classifiers on 11th generation Intel Core i5-1135G7 @ 2.40GHz 1.38 GHz 64-bit processor with 8 GB RAM.

We trained a final set of classifiers using BERT, a deep learning model that uses context to interpret text that was originally developed for use in general language-based inference tasks.^10^ BERT and related algorithms hold state-of-the-art status on a variety of NLP tasks. However, they can require large training data sets and expensive hardware, and can be complex to modify and deploy for specific use cases.^24^ In addition to ‘BERT-base’ we selected two additional BERT-related models, BioBERT^25^ and ClinicalBERT^26^. Since the original publication of BERT, several customizations have been published for use in biomedical language inference. BioBERT is one such biomedical adaptation that was generated by further training BERT using scientific abstracts and full-text articles in PubMed and PubMed Central respectively.^25^ Similarly, ClinicalBERT was trained using clinical text from MIMIC-III corpus.^27^^,^^28^ These models were fine-tuned on the training data in a NVIDIA Tesla T4 GPU environment with 16 GB RAM.

We used each of these methods to separately train a binary classifier (to predict which procedure reports were Mayo scorable) and an ordinal classifier (to predict the correct Mayo endoscopic subscore for scorable reports). Due to the presence of class imbalance, we also developed null classifiers that predict the dominant class for each task.

We used 5-fold cross-validation to select the best performing sklearn and autoML-based classifiers. All classifiers were evaluated on a 20% held-out test setstratified by score, annotator, and site. Accuracy was used as the primary criterion for classifier selection. For ordinal prediction tasks, accuracy and other binary measures of performance were computed by combining all incorrect scores into a single class.

The classifier achieving the highest accuracy was subjected to additional evaluations of generalizability and learning efficiency. To assess generalizability, we retrained the binary classifier on the data from UCSF alone and evaluated it on data from ZSFG. There were insufficient reports to adequately assess this for the ordinal prediction task due to the multiplicity of classes and class imbalance.

**Calculation of Environmental Impact**

The public ML CO2 impact calculator was utilized for estimating the environmental impact of our models. We utilized the default carbon efficiency of 0.432 kg equivalents of CO2. One graphics processing unit (NVIDIA Tesla v100) utilizes 300 watts while one central processing unit (Intel core i7) requires 100 watts.

**Mayo Endoscopic Scoring Guidance From FDA and *Keywords Used in Rule Based Classifier***

| **Mayo Score** | **Appearance of Mucosa** |
| --- | --- |
| 0 | *Normal* appearance |
| 1 | Mild*erythema*, mild *decreased vascular pattern*, no friability |
| 2 | *Marked erythema*, *absent vascular pattern*, *friability*, *erosions* |
| 3 | *Spontaneous bleeding*, *ulcer*ation |

**Annotation Protocol**

**Step 1: Exclude reports that should not be classified by the Mayo endoscopic subscore**

- Patients with a proctocolectomy (total abdominal colectomy is acceptable, to enable scoring of the Hartmann’s pouch)
- Procedures where no mucosa was visualized due to a poor bowel preparation
- Patients who are clearly diagnosed with Crohn’s disease
- E.g. Crohn’s appears in the impressions section, or it appears in the indications section only and the findings do not seem to question the diagnosis (rare)
- If the patient carries a diagnosis of UC but has CD-like features (stricture, fistulae, aphthae) and impressions section does not clearly indicate a revised diagnosis of CD, then do not exclude this
- If the patient carries a diagnosis of ‘IBD’, score as if the patient has a working diagnosis of UC

**Step 2: Classify relevant reports according to the Mayo endoscopic subscore**

Score according to the most severely affected segment of the visualized colorectum according to the following descriptors:

0: ‘normal’, ‘quiescent’, ‘scar’ without other descriptors consistent with other classes

1: ‘erythema’, ‘decreased vascular pattern’, ‘granularity’, ‘aphthous ulcer’, ‘aphthae’, ‘mild’

2: ‘friability’, marked or extensive ‘erythema’, ‘loss of (absent) vascularity’, ‘erosions’, ‘moderate’

3: spontaneous ‘bleeding’, ‘ulcer/ulcerated’, ‘severe’

Edge cases:

- Poor bowel prep: Only score the segments that are seen
- Report indicates ‘moderate to severe’ but without severe descriptors
- Classify as a 3
- Rationale: More consistent with scoring according to the worst affected segment
- Clinician assignment of a Mayo score is discordant with other descriptors(E.g. report describes the severity as mild but indicates the presence of a ulcer)
- Classify according to the most severe descriptor(e.g. “severe colitis” with erosions is a 3, “mild” colitis with an “ulcer” is a 3)
- Incomplete colonoscopy, sigmoidoscopy, proctoscopy (e.g. hartmann’s pouch):
- If the exam was intentionally halted due to distal disease severity, score as a 3
- Otherwise, only score the segments that are seen
- IBD patients with:
- Stricture: ignore from the standpoint of assigning a Mayo score
- Aphthous ulcers: score as consistent with a 1
- Fistulas: ignore from the standpoint of assigning a Mayo score
- Rationale: hard to distinguish acute from chronic
- Granular mucosa: score as consistent with a 1
- “Deep” or “fissuring” ulcer: score as a 3
- “Shallow ulcer”: score as a 3
- “Deep erosion”: score as a 2
- Scar:
- If described as associated with loss of vascularity, score as at least a 1
- Rationale: the accuracy of the endoscopic diagnosis of a chronic inflammation without activity (e.g. scar) is unknown and anecdotally, can be discordant with histology
- Otherwise, ignore
- “Mucosal healing”
- Ignore and look for other descriptors to classify the severity
- “No active inflammation”
- Ignore and look for other descriptors to classify the severity
- Patients without a clear diagnosis of IBD (e.g. could be a screening colonoscopy, or a diagnostic exam)
- So long as no exclusion criteria are met (step 1 above), assign a score per the general rules (e.g a clinical diagnosis of UC might subsequently be assigned)
- Patients with possible SCAD (erythematous mucosa): score as consistent with a 1
